# Socioeconomic inequalities in co-morbidity of overweight, obesity and mental ill-health from adolescence to mid-adulthood in two national birth cohort studies

**DOI:** 10.1101/2020.10.21.20215764

**Authors:** Amal R. Khanolkar, Praveetha Patalay

**Author notes:** Corresponding Author: Amal R. Khanolkar.

## Abstract

**Aim:** To examine socioeconomic inequalities in risk of comorbidity between overweight (including obesity) and mental ill-health in two national cohorts. We investigated independent effects of childhood and adulthood socioeconomic disadvantage on comorbidity from childhood to mid-adulthood, and differences by sex and cohort.

**Methods:** Data were from 1958 National Child Development Study (NCDS58) and 1970 British Cohort Study (BCS70) [total N=30,868, 51% males] assessed at ages 10, 16, 23, 34 and 42 years. Socioeconomic indicators included childhood and adulthood social class and educational level. Risk for i. having healthy BMI and mental ill-health, ii. overweight and good mental health, and iii. overweight and mental ill-health was analysed using multinomial logistic regression.

**Results:** Socioeconomic disadvantage was consistently associated with greater risk for overweight-mental ill-health comorbidity at all ages (RRR 1.43, 2.04, 2.38, 1.64 and 1.71 at ages 10, 16, 23, 34 and 42 respectively for unskilled/skilled vs. professional/managerial class). The observed inequalities in co-morbidity were greater than that observed for either condition alone (overweight; RRR 1.39 and 1.25, mental ill-health; 1.36 and 1.22 at ages 16 and 42 respectively, for unskilled/skilled vs. professional/managerial class). In adulthood, childhood and adulthood socioeconomic disadvantage were independently associated with comorbid overweight-mental ill-health, with a clear inverse gradient between educational level and risk for comorbidity; no education, RRR 6.11 (95% CI 4.31-8.65) at age 34 and 4.42 (3.28-5.96) at age 42 compared to university education. There were no differences observed in the extent of inequalities by sex and differences between cohorts were limited.

**Conclusions:** Socioeconomic disadvantage in childhood and adulthood are consistently and independently associated with greater risk for mental ill-health and overweight separately, and even greater inequalities in the risk for comorbidity between both conditions through the lifecourse. These findings are significant given the increasing global prevalence of obesity and mental ill-health, and their implications for lifelong health and mortality.

## Introduction

Obesity and mental ill-health are often childhood-onset, are chronic with a strong propensity to track across the lifecourse, and are associated with increased risk for a wide range of health outcomes and mortality(1, 2). Both conditions have increased in prevalence globally and contribute significantly to global disease burden(3-5) Additionally, separately both obesity and mental ill-health are more common among socioeconomically disadvantaged individuals (6-9). There is substantial evidence for comorbidity between being obese and mental ill-health (like depression and anxiety)(10-14).

Given the strong socioeconomic patterning in obesity and mental ill-health across the lifecourse, it is possible that socioeconomically disadvantaged individuals are at higher risk of comorbidity between the two conditions (above and beyond the risk of either on its own), and that this widens with age. Comorbidity is potentially more common in younger generations who experience higher rates of obesity and mental ill-health earlier in life(15, 16). This is substantiated by evidence showing inequalities in childhood obesity have widened across generations in the last few decades, which might result in inequalities in co-morbidities not being the same across generations(17, 18). There is also some limited evidence that women are at higher risk for comorbid obesity and mental ill-health, but there is little known on whether disadvantaged women are more likely to experience higher rates of comorbidity(19, 20). Disadvantaged socioeconomic circumstances in childhood can have long-lasting impacts on health across the life course including obesity and mental ill-health(21, 22). Analysing the independent effects of childhood and adulthood socioeconomic circumstances on obesity and mental ill-health comorbidity could guide shaping future public health interventions.

The limited evidence on higher rates of obesity and mental ill-health comorbidity in socioeconomically disadvantaged groups are from cross-sectional studies in adulthood (2, 14). No study has comprehensively examined socioeconomic inequalities between the two conditions using multiple socioeconomic indicators, and if it changes across the lifecourse. If socioeconomic inequalities in comorbidity between obesity and mental ill-health exist, then the health, economic consequences, and additional burden on the public healthcare systems would be much larger than that posed by either condition alone. Additionally, comorbidity between the two conditions is likely to increase with age, as obesity increases across adulthood but also increases in either condition can lead to a rise in the other over time (negative feedback loop)(13), which highlights the importance to study comorbidity in the same individuals across the lifecourse. This could also provide further explanation for existing and persistent inequalities in morbidity and mortality observed globally.

This study investigated childhood and adulthood socioeconomic inequalities in comorbidity between obesity and mental ill-health using longitudinal data from two large nationally representative birth cohorts initiated 12 years apart. This enabled examining change in socioeconomic inequalities across time (from childhood to mid-adulthood), and additionally to study differences in socioeconomic patterning by sex and cohort.

## Methods

### Data, setting and study population

For this study, we utilised data from two ongoing national birth cohort studies: the 1958 National Child Development Study (NCDS58) and the 1970 British Cohort Study (BCS70) (23, 24). Both studies were initiated as birth surveys and are prospective and longitudinal; the NCDS58 follows the lives of an initial 17,415 people born in a single week in March 1958 and the BCS70 follows 17,198 people born in a single week in April 1970 in England, Scotland and Wales. More information on the two cohorts can be found at: https://cls.ucl.ac.uk/cls-studies/. Data for this study was drawn from 5 waves at which BMI and mental health symptoms were both assessed; ages 11, 16, 23, 33 and 42 in NCDS58 and ages 10, 16, 26, 34 and 42 in BCS70 (here after ages 10, 16, 23, 34 and 42 for simplicity). Eligible participants were those with a minimum of one datapoint on BMI or mental health recorded at any one of the 5 waves, yielding a final study sample of N=30,868 (NCDS58, N=16,464 and BCS70, N=14,404, 95% and 84% of the initial cohorts respectively).

#### Mental health

Poor mental health is based on measures of psychological distress which assesses symptoms of common mental health disorders such as anxiety and depression. Psychological distress was assessed by questionnaires answered by participants (or their parents during childhood) from both cohorts. The Rutter internalising scale consists of five items describing depressive and anxiety symptoms measured consistently in both childhood sweeps (ages 10 and 16) and in both cohorts which can be combined to give an index of emotional difficulties in children (for example ‘*Often worried, worries about many things’*). Parents were asked to indicate whether each description ‘does not apply’, ‘applies somewhat’ or ‘definitely applies’ to a child.

The 9-item Malaise Inventory (a set of ‘yes-no’ self-completion questions like ‘*Do you often feel miserable or depressed?’*) measures levels of psychological distress including symptoms of both anxiety and depression in adults and measured consistently in all adulthood sweeps in both cohorts (ages 23, 34 and 42) is used here. Items from the Rutter scale and Malaise inventory are listed in Supplementary Table 1.

**Table 1.** Descriptive statistics of variables of interest in 30,868 participants from the 1958 National Child Development Study and the 1970 British Cohort Study. Numbers are means or percentages.

|  | Age 11/10 | Age 16 | Age 23/26 | Age 33/34 | Age 42 |
| --- | --- | --- | --- | --- | --- |
|  | Mean or % (95% CI) | Mean or % (95% CI) | Mean or % (95% CI) | Mean or % (95% CI) | Mean or % (95% CI) |
| **Full study sample, N=30,868** |  |  |  |  |  |
| Mental health | 2.78 (2.75-2.81) | 2.04 (2.01-2.06) | 1.53 (1.5-1.56) | 1.36 (1.33-1.38) | 1.74 (1.71-1.77) |
| High psych distress (%) | 20 (19.2-20.2) | 18 (17.6-18.6) | 11.5 (11-11.9) | 10.4 (10-10.8) | 15 (14.3-15.3) |
| BMI (kg/m<sup>2</sup>) | 17.2 (17.1-17.2) | 20.9 (20.8-20.92) | 23.2 (23.1-23.3) | 25.5 (25.4-25.5) | 26.4 (26.3-26.5) |
| Overweight or Obesity (%) | 9 (8.7-9.4) | 12.3 (11.9-12.8) | 25.6 (25.1-26.2) | 49 (48.3-49.6) | 57.5 (56.8-58) |
| **Outcome of interest (%)** |  |  |  |  |  |
| Healthy BMI & good MH | 73 (72.5-74) | 71.8 (71.3-72.4) | 66 (65.5-66.7) | 46 (45.4-46.6) | 36.6 (35.9-37.2) |
| OW/OB & good MH | 7 (6.9-7.5) | 10 (9.5-10) | 22.4 (21.9-22.9) | 44 (42.9-44.2) | 48.6 (47.9-49.2) |
| Healthy BMI & poor MH | 17.8 (17.4-18.3) | 15.7 (15.3-16) | 8.3 (7.9-8.7) | 5 (4.8-5.3) | 6 (5.6-6.3) |
| OW/OB & poor MH | 1.8 (1.6-1.9) | 2.4 (2.1-2.5) | 3.2 (2.9-3.4) | 5.3 (5-5.6) | 8.9 (8.4-9.2) |
| **NCD58 only, N=16,464** |  |  |  |  |  |
| Mental health | 3.26 (3.23-3.29) | 1.96 (1.93-1.99) | 1.27 (1.24-1.30) | 1.04 (1.01-1.06) | 1.56 (1.52-1.59) |
| High psych distress (%) | 23.4 (22.7-24.1) | 16.7 (16-17.3) | 8.8 (8.3-9.2) | 6.6 (6.1-7) | 12.2 (11.7-12.8) |
| BMI (kg/m<sup>2</sup>) | 17.5 (17.44-17.53) | 20.68 (20.62-20.73) | 22.65 (22.6-22.71) | 25.03 (24.96-25.11) | 25.96 (25.88-26.04) |
| Overweight or Obesity (%) | 9.6 (9-10) | 11 (10.4-11.5) | 18.7 (18-19.4) | 45 (44.1-45.9) | 54.1 (53.2-54.9) |
| **Outcome of interest (%)** |  |  |  |  |  |
| Healthy BMI & good MH | 69.2 (68.4-70) | 74.2 (73.4-75) | 74.2 (73.4-75) | 51.3 (50.4-52.2) | 40.3 (39.4-41.1) |
| OW/OB & good MH | 7.4 (7-7.8) | 9.1 (8.6-9.6) | 17 (16.3-17.6) | 42 (41.2-43) | 47.5 (46.6-48.4) |
| Healthy BMI & poor MH | 21.3 (20.6-21.9) | 14.9 (14.2-15.5) | 7 (6.6-7.4) | 3.7 (3.4-4) | 5.6 (5.3-6.1) |
| OW/OB & poor MH | 2.2 (1.9-2.4) | 1.8 (1.6-2) | 1.8 (1.5-2) | 3 (2.6-3.2) | 6.6 (6.1-6.9) |
| **BCS70 only, N=14,404** |  |  |  |  |  |
| Mental health | 2.23 (2.19-2.27) | 2.12 (2.08-2.16) | 1.83 (1.80-1.86) | 1.73 (1.70-1.77) | 1.95 (1.90-1.99) |
| High psych distress (%) | 15.4 (14.8-16) | 20 (19.1-20.6) | 14.6 (13.9-15.3) | 14.7 (14-15.3) | 18 (16.9-18.6) |
| BMI (kg/m<sup>2</sup>) | 16.88 (16.84-16.92) | 21.11 (21.05-21.17) | 23.87 (23.78-23.95) | 25.95 (25.85-16.04) | 26.98 (26.88-27.09) |
| Overweight or Obesity (%) | 8.5 (7.9-8.9) | 14 (13.2-14.6) | 33.5 (32.4-34.4) | 53.4 (52.4-54.4) | 61.3 (60.4-62.3) |
| **Outcome of interest (%)** |  |  |  |  |  |
| Healthy BMI & good MH | 77.5 (76.8-78.2) | 69.2 (68.4-70) | 56.8 (55.7-57.8) | 40 (39.1-41) | 32.4 (31.4-33.3) |
| OW/OB & good MH | 7 (6.6-7.5) | 11 (10.3-11.6) | 28.7 (27.7-29.6) | 45.3 (44.3-46.4) | 50 (48.8-50.1) |
| Healthy BMI & poor MH | 14 (13.4-14.6) | 16.8 (16.1-17.6) | 9.7 (9.2-10.4) | 6.6 (6.1-7.1) | 6.3 (5.7-6.8) |
| OW/OB & poor MH | 1.4 (1.3-1.5) | 3 (2.6-3.3) | 4.8 (4.3-5.2) | 8 (7.5-8.6) | 11.5 (10.8-12.2) |

Overall scores were calculated separately for the Rutter and Malaise scales at each sweep by aggregating the item responses to create a total score used in analysis (with higher scores indicating higher levels of internalising symptoms and psychological distress). These overall scores were dichotomized (score ≥4) for the Malaise-9 to identify individuals with and without high levels of psychological distress. There is no established cut-off point for dichotomising the Rutter scale. Based on previous research we chose the 85^th^ percentile as the cut-off point (which corresponds to scores ≥5 and ≥4 for ages 10 and 16 respectively).

#### Body mass index

BMI [weight (kg)/height (m)^2^] was calculated from objectively assessed height and weight (or self-reported when objective measures were missing at ages 26, 34 and 42 in BCS70 and ages 23 and 42 in NCDS58). Overweight and obesity in childhood were defined using the age- and sex-specific cut-offs proposed by International Obesity Task Force (IOTF)(25). The WHO criteria of 25-29.99kg/m^2^ and ≥30kg/m^2^ were used for calculating overweight and obesity respectively in adulthood. In all analyses the overweight and obese groups were combined (referred as overweight hereafter) and compared to the healthy weight group (which also included the underweight group [<18.5kg/m^2^]).

#### Predictors of interest

Our predictors of interest included social class (father’s social class measured at age 10 and participant’s own social class measured at age 42), and highest educational level (ascertained at age 33). To ensure cross-cohort comparability, the UK Registrar General’s 1990 version was used to classify both parental and participant’s own social class. The original 6 categories were grouped as follows for analysis: 1. Professional & managerial (reference category), 2. Non-manual, 3. Manual and 4. Partly skilled & unskilled. Educational levels are based on the highest UK National Vocational Qualifications (NVQ) level (listed in Supplemental Table 3) and included six categories: no education, NVQ level 1 which represents General Certificate of Secondary Education grade D-G or lower; NVQ level 2, General Certificate of Secondary Education grades A*-C and equivalent qualifications; NVQ level 3, A-levels and equivalent; NVQ level 4, earning a degree; and NVQ level 5, university degree or diploma (reference category). Education was harmonised across both cohorts ensuring comparability.

#### Comorbidity indicator (study outcome)

The outcome of interest was created separately at each age by combining the dichotomised BMI and mental health variables resulting in a variable with four categories: 1. healthy BMI and good mental health (reference group [↑BMI-MH↑], 2. healthy BMI and poor mental health [↑BMI-MH↓], 3. overweight or obesity and good mental health [↓BMI-MH↑] and 4. overweight or obesity and poor mental health [↓BMI-MH↓].

### Data Analysis

Initial analysis included descriptive statistics of BMI and mental health at each age including means, variances and distributions for continuous measures and prevalence of overweight/obesity and poor mental health (high psychological distress) in the entire study sample and by cohort and sex. We also estimated mean differences in BMI and mental health by socioeconomic indictors in the full study sample and by cohort.

We used multivariable multinomial logistic regression to estimate relative risk ratios (RRR) of i. ↑BMI-MH↓ ii. ↓BMI-MH↑ and iii. ↓BMI-MH↓ with healthy BMI and good mental health as the baseline, comparing more socioeconomically disadvantaged participants with the most advantaged. The multinomial logistic regression model calculates the relative risk ratio (RRR), which is the ratio of two relative risks and is interpreted for a unit change in the predictor variables.

The following models were run separately at the different age sweeps in the full study sample to assess RRR for the different categories of the outcome variable across the categories of socioeconomic predictors: ages 10, 16 and 23; associations with childhood social class, age 34; associations with childhood social class and educational level, age 42; associations with childhood social class, educational level and adulthood social class. All models were adjusted for gender and cohort.

We also examined associations between childhood social class and risk for the different categories of the comorbidity indicator at each age (to help understand the independent effect of early life social class on comorbidity in both childhood and adulthood). These models were only adjusted for gender.

We tested whether the above associations between the three socioeconomic indicators and the outcome variable differed by 1. cohort and 2. gender by including interactions terms between each socioeconomic indicator and cohort or gender. Interactions tests between gender and socioeconomic indictors were not significant and results are not presented.

Missing data in the five sweeps in both cohorts was accounted for using multiple imputation assuming data missing at random (and including 25 imputations). All analyses were run in Stata 15 (College Station, TX, USA).

## Results

Table 1 presents descriptive statistics for BMI and mental health, and the outcome variable (comorbidity of overweight and mental ill-health) across the five age sweeps. On average, mean BMI increased from 17.2kg/m^2^ at age 10 to 26.4kg/m^2^ at age 42. From age 16 onwards, the younger BCS70 cohort higher mean BMI and higher proportions of participants being overweight compared to the NCDS58 cohort (for example, 19% vs. 33% at age 23 and 54% vs. 61% at age 42 being overweight in the NCDS58 and BCS70 cohorts respectively). Across adulthood, the younger BCS70 participants reported higher proportions of mental ill-health (for example 8.8 vs 14.6% at age 23 and 12.2 vs. 18% at age 42 in the NCDS58 and BCS70 cohorts respectively).

The proportion of participants with ↑BMI-MH↑ decreased progressively from 73% at age 10 to 37% at age 42. The largest increase was observed in the ↓BMI-MH↑ group (7% at age 10 to 49% at age 42). The ↓BMI-MH↓ group increased from 2% at age 10 to 9% at age 42. Across adulthood the proportion of participants with ↓BMI-MH↓ comorbidity was higher in the younger BCS70 cohort (for example, 2 vs. 5% at age 23 and 7 vs.12% at age 42 in NCDS58 and BCS70 cohorts respectively).

A higher proportion of BCS70 participants were in the most advantaged social class group (Table 2, 47 vs. 40% for adulthood social class and 27 vs. 23% for childhood social class in BCS70 and NCDS58 respectively).

**Table 2.** Distribution of 30,868 participants from the 1958 National Child Development Study and the 1970 British Cohort Study by gender and socioeconomic indicators of interest. Numbers are percentages (95% CI).

| **Covariates** | **Full study sample**<br>**N=30,868** | **NCDS58**<br>**N=16,464** | **BCS70**<br>**N=14,404** |
| --- | --- | --- | --- |
| **Sex** |  |  |  |
| Males | 51.4 (50.8-51.9) | 51.3 (50.6-52.1) | 51.4 (50.6-52.2) |
| Females | 48.6 (48.1-49.2) | 48.7 (47.9-49.4) | 48.6 (47.7-49.4) |
| **Childhood social class** |  |  |  |
| Professional & Managerial | 24.6 (24.1-25.1) | 22.7 (22.1-23.4) | 26.7 (25.9-27.4) |
| Non-manual | 10.4 (10-10.7) | 10.2 (9.7-19.6) | 10.6 (10.1-11.1) |
| Manual | 42.2 (41.6-42.7) | 42.9 (42.2-43.7) | 41.2 (40.4-42.1) |
| Partly skilled & Unskilled | 22.9 (22.4-23.4) | 24.1 (23.5-24.8) | 21.5 (20.8-22.1) |
| **Adulthood social class** |  |  |  |
| Professional & Managerial | 43.6 (42.8-44.3) | 40.1 (39.2-41) | 47.5 (46.4-48.7) |
| Non-manual | 20.2 (19.6-20.8) | 21.6 (20.8-22.4) | 18.6 (17.7-19.4) |
| Manual | 19.9 (19.3-20.5) | 21 (20.2-21.8) | 18.5 (17.6-19.4) |
| Partly skilled & Unskilled | 16.4 (15.7-17) | 17.3 (16.5-18) | 15.4 (14.4-16.4) |
| **Highest educational level** |  |  |  |
| None | 12 (11.4-12.4) | 13.3 (12.7-13.9) | 10.3 (9.6-10.9) |
| Nvq1 level (Secondary education, lower grades) | 10.7 (10.3-1.1) | 12.8 (12.2-13.4) | 8.4 (7.8-8.9) |
| Nvq2 level (Secondary education, higher grades) | 32 (31.3-32.6) | 34 (33.1-34.9) | 29.7 (28.7-30.7) |
| Nvq3 level (School leaving certificate, A-levels) | 14.2 (13.7-14.7) | 13.9 (13.3-14.6) | 14.5 (13.8-15.3) |
| Nvq4 level (Earning a degree) | 21.8 (21.3-22.4) | 13.8 (13.2-14.5) | 31 (30-31.9) |
| Nvq5 level (University degree or diploma) | 9.3 (8.9-9.6) | 12.1 (11.5-12.7) | 6.1 (5.7-6.6) |

### Inequalities in BMI and mental health over time

Supplemental Tables 2A and 2B show the variation in mean BMI and mental health by socioeconomic indicators at each age. In childhood, disadvantaged social class groups had higher levels of mental ill-health compared to the most advantaged. Similarly, differences in mean BMI were more visible at age 16 compared to age 10 and larger in BCS70 compared to NCDS58 (mean BMI 20.5 vs. 20.8kg/m^2^ in NCDS58 and 20.9 vs. 21.4kg/m^2^ in BCS70 for professional/managerial vs. partly skilled/unskilled respectively).

Differences in mean BMI and mental health between the most and least advantaged got wider, more consistent with age and were observed with all three socioeconomic indicators. The largest differences in mean BMI and mental health were observed with educational level (with a clear inverse gradient) in both cohorts. These differences were larger in the BCS70 cohort (for example, the mean difference in BMI for no education compared to having a degree was 1.8kg/m^2^ in NCDS58 and 2.5 kg/m^2^ in BCS70 at age 33, which increased to 2.3 kg/m^2^ in NCDS58 and 3kg/m^2^ in BCS70 at age 42). Mean differences in mental health between the most and least advantaged groups decreased marginally across adulthood.

### Socioeconomic inequalities in risk for comorbidity between overweight and mental ill-health over time

Supplemental Table 3 and Figures 1 and 2 display results from multinomial logistic regression examining risk for the different combinations of categorical BMI and mental health at each age. In general, the socioeconomically disadvantaged had higher risks for overweight and mental ill-health independently and comorbidity at all ages. In childhood, participants from more disadvantaged families had consistently higher relative risk ratios (RRR) for the three adverse groups. Compared to the most advantaged (professional/managerial), the most disadvantaged group (partly skilled/unskilled) had the highest RRR for all three combinations of BMI and mental health (for example at age 16, RRR 1.39 [95% CI 1.21-1.58] for ↓BMI-MH↑, RRR 1.36 [1.22-1.51] for ↑BMI-MH↓, and RRR 2.04 [1.54-2.72] for ↓BMI-MH↓).

**Figure 1 A, B, C.**
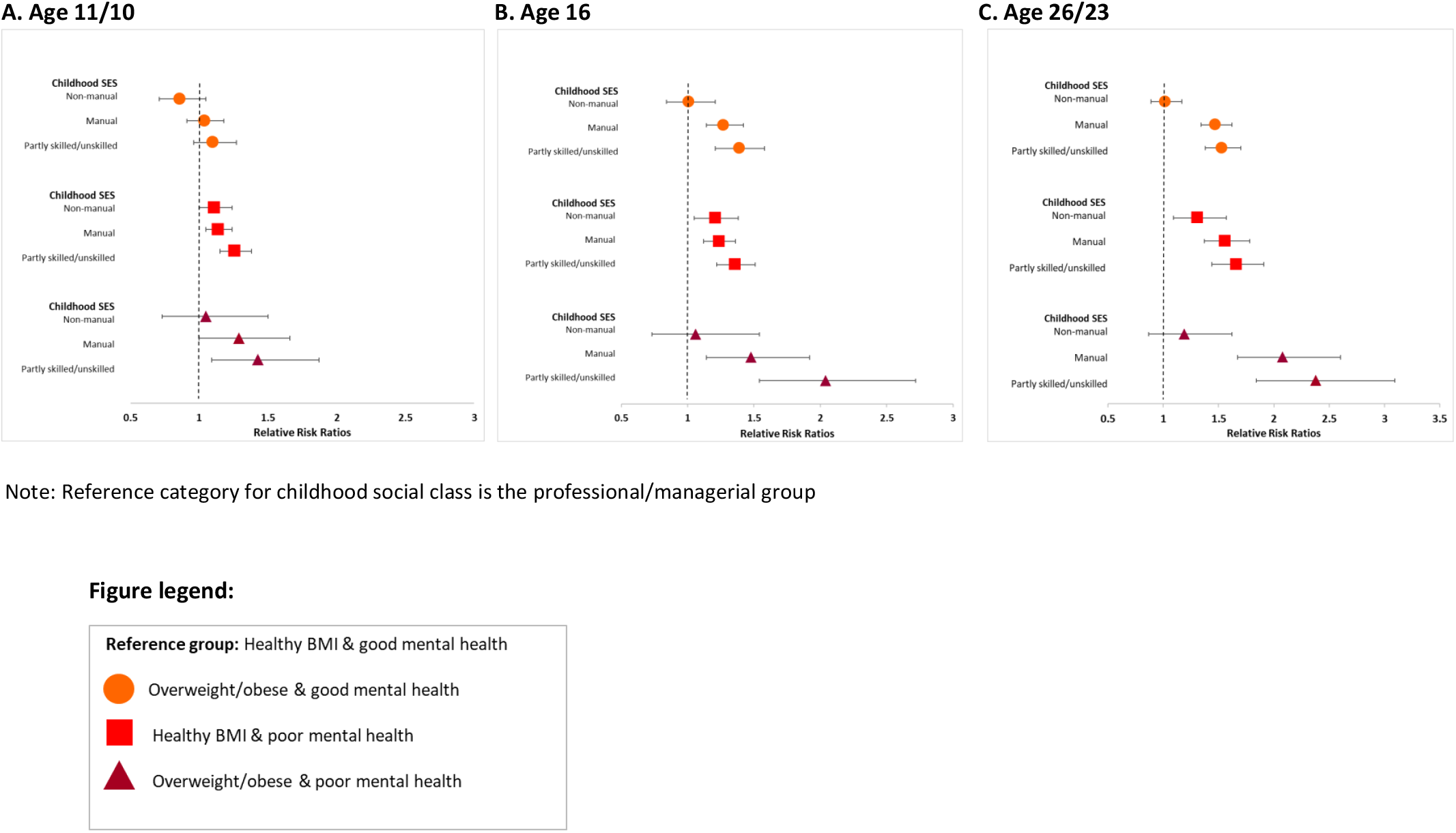
Relative risk ratios (RRR) for i. overweight or obesity and good mental health, ii. Healthy BMI and poor mental health and iii. Overweight or obesity and poor mental health in 30,868 participants from the 1958 National Child Development Study and the 1970 British Cohort Study. Healthy BMI and good mental health is the reference category

**Figure 2 A & B.**
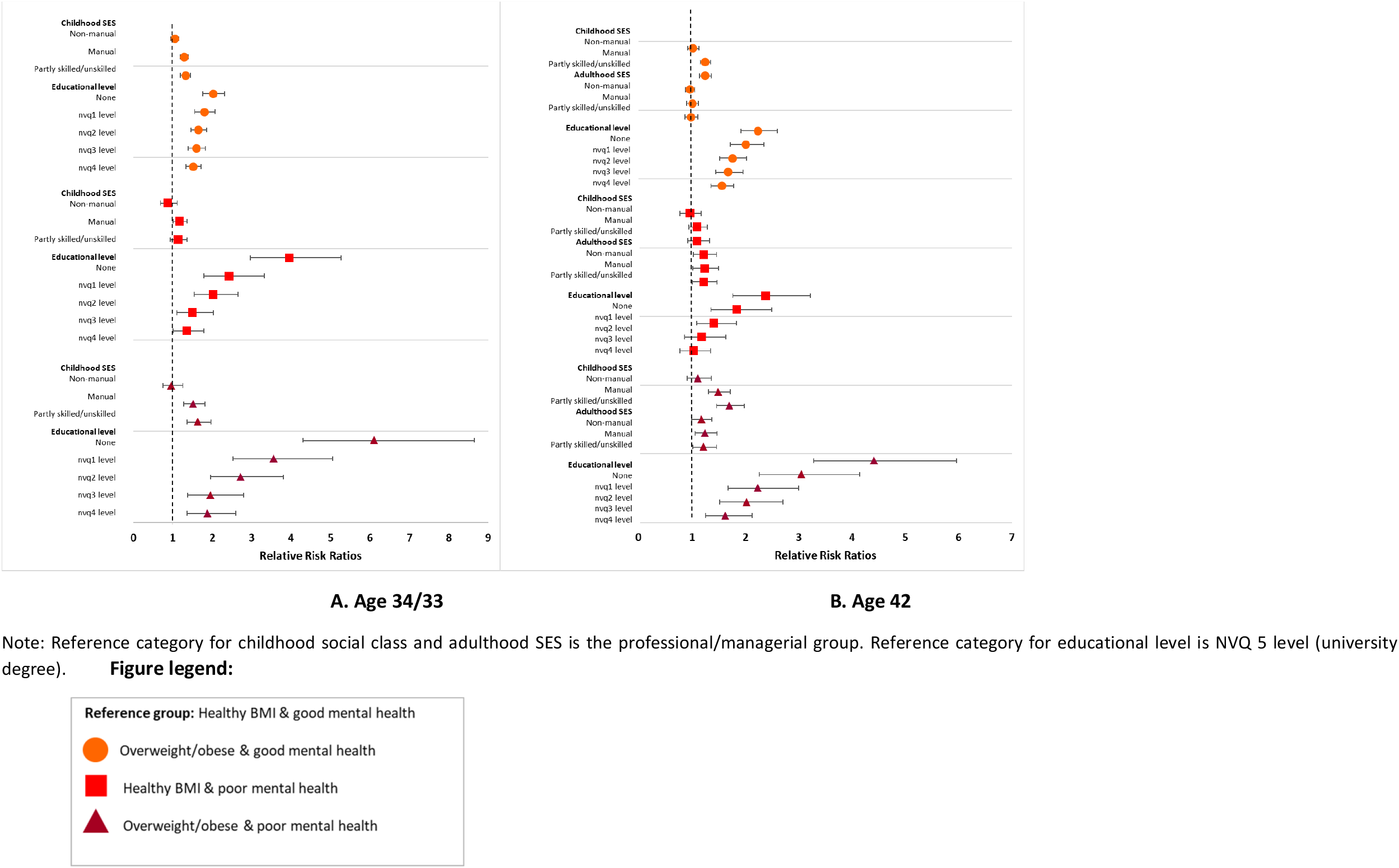
Relative risk ratios (RRR) for i. overweight or obesity and good mental health, ii. Healthy BMI and poor mental health and iii. Overweight or obesity and poor mental health in 30,868 participants from the 1958 National Child Development Study and the 1970 British Cohort Study. Healthy BMI and good mental health is the reference category

The associations between childhood social class and risk for the three adverse combinations of BMI and mental health were larger in effect size and observed consistently for both manual and partly skilled/unskilled groups at age 23 (compared to associations seen at ages 10 and 16).

There was a clear inverse gradient between educational level and risk for the three adverse combinations of BMI and mental health at ages 34 and 42. Compared to those with a university degree, all other educational categories had higher risk for the three adverse combinations of BMI and mental health, with the risk increasing as educational levels decreased, even after mutual adjustment for other socioeconomic indicators.

A disadvantaged childhood social class was independently associated with increased risk of ↓BMI-MH↓ comorbidity in adulthood (adjusted RRR 2.38 [1.84-3.09] at age 23 and 1.71[1.46-1.99] at age 42 for partly skilled/unskilled group).

Compared to NCDS58 participants, BCS70 participants had an increased risk for all three adverse combinations of BMI and mental health with the largest risks for ↓BMI-MH↓ comorbidity (Supplemental Table 3, RRR 3.69 [3.13-4.35] at age 23 and 3.95 [3.47-4.5] at age 34.

Figure 3 and Supplemental Table 5 display results from multinomial regression modelling examining risk of comorbidity with only childhood social class at all ages and stratified by cohort. At all ages, a disadvantaged childhood social class was associated with increased risk for all adverse groups, with RRR getting larger with age. The largest risk was observed for ↓BMI-MH↓ comorbidity at all ages and in both cohorts (for example, RRR 1.71 [1.12-2.61] and 2.33 [1.63-3.32] at age 16 and RRR 2.1 [1.70-2.60] and 2.33 [1.87-2.89] at age 42 for the partly skilled/unskilled group in NCDS58 and BCS70 respectively). While BCS70 participants had higher RRR for ↓BMI-MH↓ comorbidity in childhood compared to NCDS70, the reverse was observed at ages 23 and 34. However, confidence intervals for cohort-specific RRR were largely overlapping.

**Figure 3.**
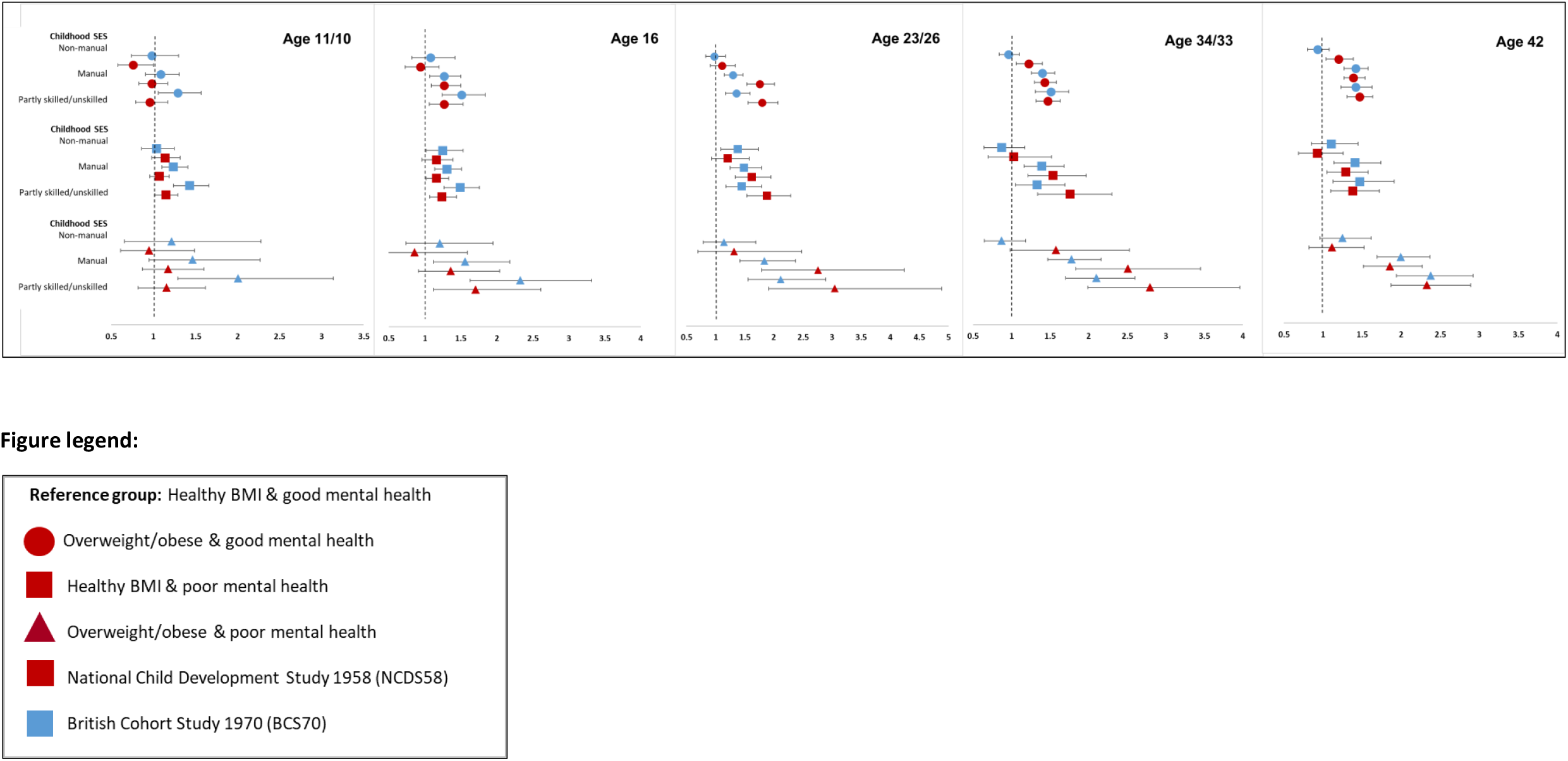
Relative risk ratios (RRR) between childhood social class and risk for i. overweight or obesity and good mental health, ii. Healthy BMI and poor mental health and iii. Overweight or obesity and poor mental health in 30,868 participants from the 1958 National Child Development Study and the 1970 British Cohort Study. Healthy BMI and good mental health is the reference category

### Differences in socioeconomic inequalities in comorbidity by cohort and gender

Risk for the three adverse combinations of comorbidity between the most and least advantaged differed between the two cohorts at all ages except age 16 (tests for interaction; age 11/10, p<0.05, ages 23, 34/33 and 42, p<0.001) (Supplemental Table 4 and Supplemental Figures 2). At age 11/10, participants in the younger BCS70 cohort had higher risk for all three adverse combinations compared to the NCDS58 cohort (Supplemental Figure 2A). At ages 34/33 and 42, NCDS58 participants with no education had higher risk of ↑BMI-MH↓ and ↓BMI-MH↓ comorbidity compared to BCS70 participants (Supplemental Figures 2C and 2D).

Interactions tests between gender and socioeconomic indictors indicated that the socioeconomic patterns observed are similar across males and females.

## Discussion

This study found 1. Consistent associations between socioeconomic disadvantage and increased risk of co-morbid overweight and mental ill-health from childhood into mid-adulthood, 2. While socioeconomic disadvantage was associated with increased risks of either overweight or mental ill-health in childhood and adulthood, the largest risks for inequalities were observed for their comorbidity. 3. Greater levels of socioeconomic disadvantage were associated with increasing risks of comorbidity, with inverse gradients observed with social class and educational level, 4. Disadvantaged childhood social class was associated with increased risk of comorbidity in adulthood even after accounting for adulthood socioeconomic circumstances. 5. Differences by cohort and sex in the extent of socioeconomic inequalities in risk for comorbidity were limited.

This study benefits from a large sample size drawn from two nationally representative birth cohorts with multiple BMI and mental health measurements. Like most longitudinal studies, both cohorts suffer from attrition but was addressed using the robust multiple-imputation technique. While BMI was self-reported at some ages in both cohorts, studies indicate that self-reported measurements do not substantially bias estimates(26). Extent of inequalities for mental health in childhood could be overestimated, as previous studies show greater socioeconomic inequalities in mental health when using parent-compared to individual-reported data(27). This study benefits from the use of childhood and adulthood socioeconomic indicators which help understand the independent contribution of both markers on comorbidity across time. Inferences on cross-cohort comparisons are strengthened by using harmonised socioeconomic indicators, BMI and mental health measures.

To our knowledge no study has examined socioeconomic inequalities in risk of co-morbid higher BMI and mental ill-health in the same individuals over time. However, socioeconomic inequalities in BMI and mental health are separately well established across the lifecourse and these inequalities have increased in younger generations(8, 15, 17). Being socioeconomically disadvantaged is not only a common risk factor for both poor mental health and obesity, it is linked to other factors on the pathway to either condition. For example, living in disadvantaged neighbourhoods is often associated with decreased access to recreational areas, green spaces and healthier food(28-30). The association between obesity and poor mental health is bidirectional (obesity leads to poor mental health and vice versa(31)) but both conditions can be a result of common risk factors more likely to impact disadvantaged groups like increased stress levels, health behaviours (poorer eating habits, inadequate exercise), social stigma, higher unemployment etc. Socioeconomically disadvantaged individuals are more likely to experience ongoing and continuing stressors which impact their health across the lifecourse leading to an accumulative effect which perpetuates health inequalities (8). Studies have indicated common biological pathways that might explain the link between obesity and depression, particularly the activation of the hypothalamic–pituitary–adrenal (HPA) axis(32). Exposure to early life stress and chronic stress over time can lead to hyperactivation of the HPA-axis resulting in high levels of cortisol associated with increased risk for both depression and obesity.

Prevalence of obesity and mental ill-health were higher in adulthood in the younger BCS70 cohort, yet cohort differences in socioeconomic inequalities in comorbidity were limited (with some indication of greater inequalities in the older NCDS58 cohort). One reason could be the shift in distribution of socioeconomic position, with the younger BCS cohort having higher number of individuals in the more socioeconomic advantaged groups. It is also possible that socioeconomic inequalities in comorbidity might manifest differently at older ages in the two cohorts which can be examined in the future. Socioeconomically disadvantaged women are more vulnerable to obesity and depression compared to men, yet our study found no sex differences in socioeconomic inequalities in comorbidity(33-36).

Health inequalities have not only persisted but widened in recent decades despite significant policies developed to tackle them. The theory of syndemics suggests that diseases cluster together producing greater impacts on health (synergism) at the population- and individual-level and is one of the mechanisms by which health inequalities develop and perpetuate in the socioeconomically disadvantaged(37, 38). Hence, any model developed to reduce inequalities associated with comorbidity will have to address not only obesity and mental ill-health but also associated risk factors and conditions that drive them while incorporating macro- and individual-level factors. A syndemic approach in reducing inequalities should include targeting common root causes of comorbid conditions at neighbourhood-level (like promoting healthier lifestyles like better and easy access to recreational spaces, affordable gyms, and healthy food options, integrated healthcare services, while accounting for local population factors like unemployment and ethnic diversity for example). Additionally, such services should be more accessible to disadvantaged populations (that is while universal in nature, they must be proportional with increased targeting towards the more disadvantaged like unemployed, low income, lone parents or ethnic-minority individuals without creating any stigma). Lastly, public health policies to reduce inequalities must have a lifecourse dimension targeting all ages as conditions like obesity and mental ill-health are more likely to occur separately and together in socioeconomically disadvantaged groups across the lifecourse as shown in this study.

In conclusion, this large study on two national birth cohorts found robust evidence for socioeconomic inequalities in risk of being overweight, having mental ill-health and comorbidity between the two conditions in individuals followed from childhood to mid-adulthood. Associations were consistent with both childhood and adulthood socioeconomic indicators and got progressively larger over time, with the most disadvantaged individuals experiencing the greatest risk. Lastly, socioeconomic disadvantage in childhood was independently associated with increased risk of comorbidity in mid-adulthood.

## Data Availability

Data for this study is from the 1958 National Child Development Study (NCDS) and 1970 the British Cohort Study (BCS). Anonymised data from both cohorts can be downloaded from the UK data service website.

## Declarations

### Availability of data and material (data transparency)

Anonymised data from the 1958 National Survey of Health and Development (NCDS) and the 1970 British Cohort Study (BCS) can be obtained free of charge from the UK Data Service (for more information: https://cls.ucl.ac.uk/data-access-training/).

### Code availability (software application or custom code)

Available upon request.

### Authors’ contributions

ARK and PP conceived the research idea. AK conducted the data preparation and data analysis. Both authors were involved in the research design, interpretation of results and writing the manuscript. Both authors read and approved the final submitted manuscript.

## Supplemental Data (Tables & Figures)

**Supplemental Table 1.**
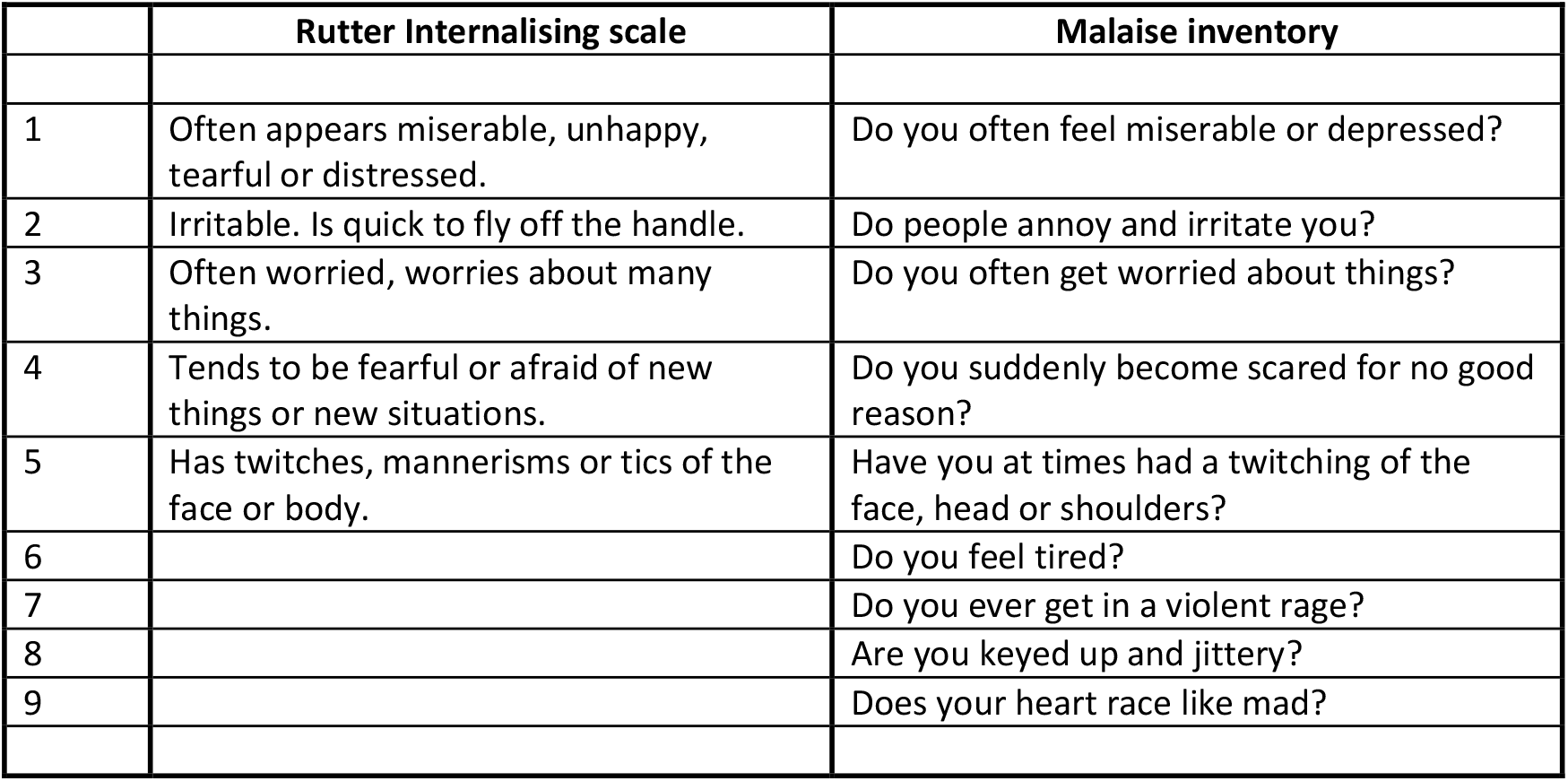
Questions that comprise the Rutter internalising scale and the Malaise inventory used to measure internalising symptoms The first five items listed are common to both scales.

**Supplemental Table 2A.**
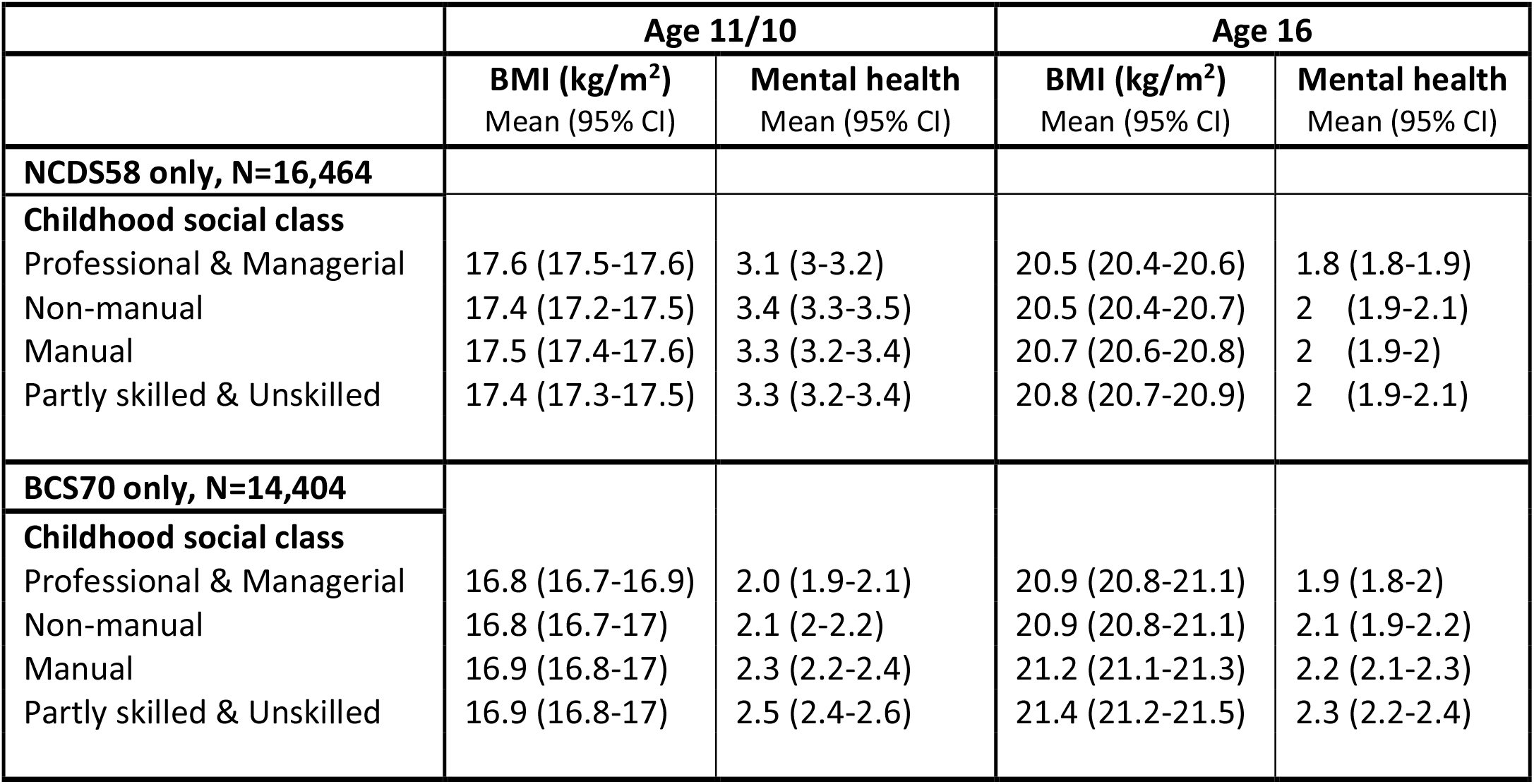
Distributions of BMI and mental health (high psychological distress) in 30 868 participants from the 1958 National Child Development Study and the 1970 British Cohort Study by socioeconomic indicators in childhood.

**Supplemental Table 2B.**
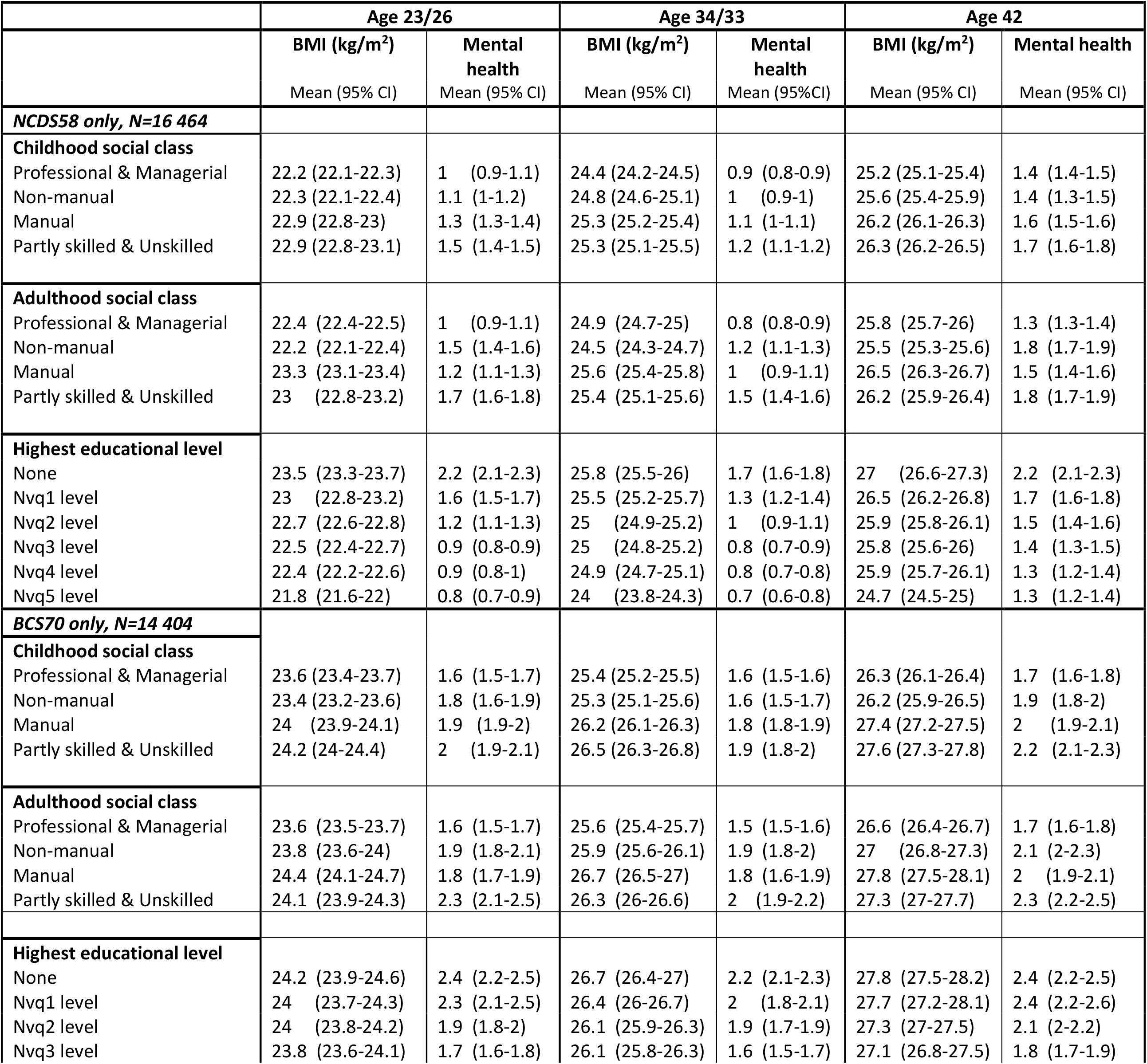
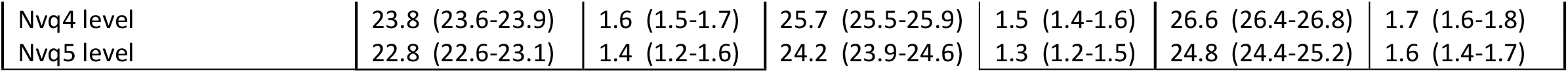
Distributions of BMI and mental health (high psychological distress) in 30 868 participants from the 1958 National Child Development Study and the 1970 British Cohort Study by socioeconomic indicators in adulthood.

**Supplemental Table 3.**
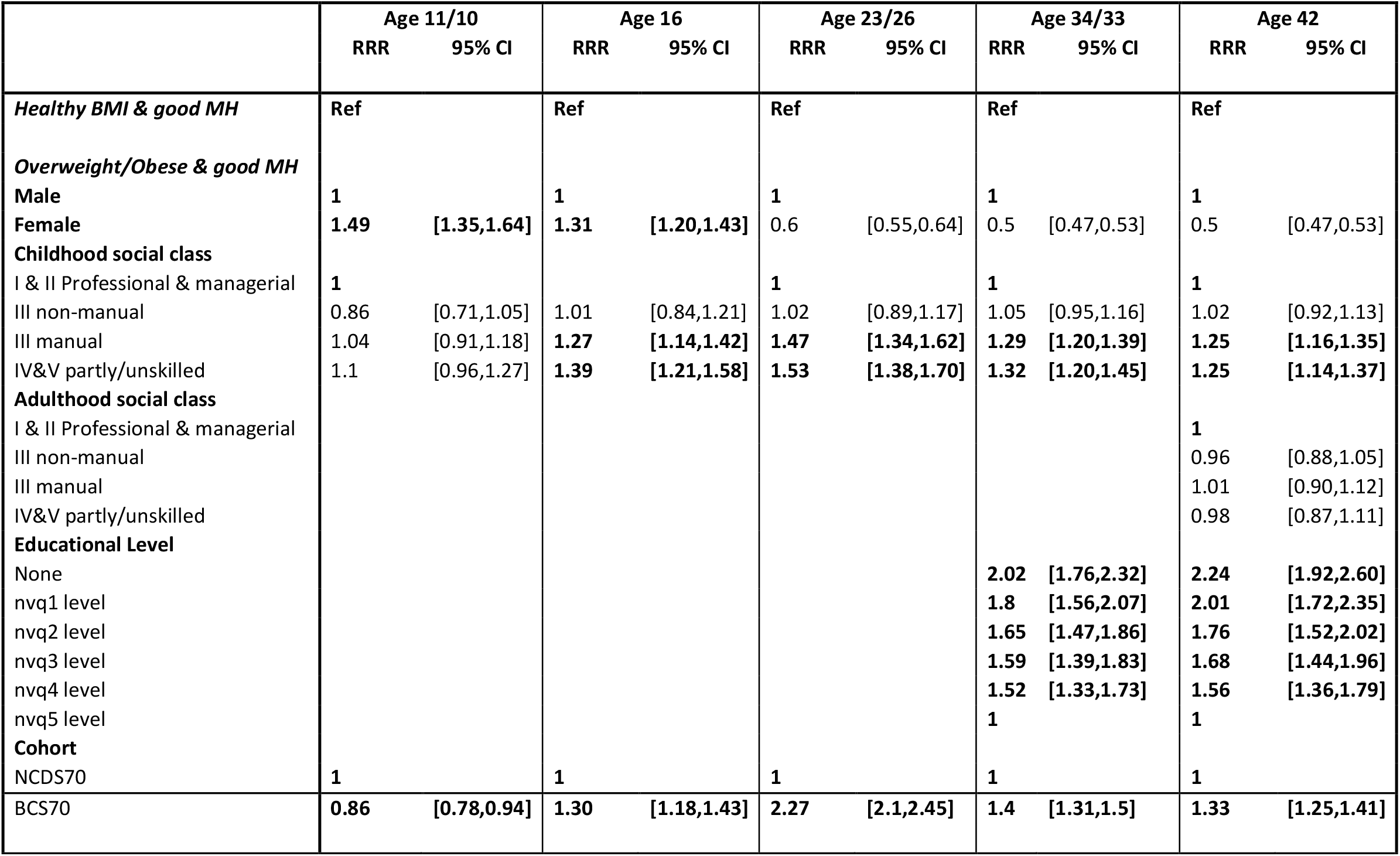
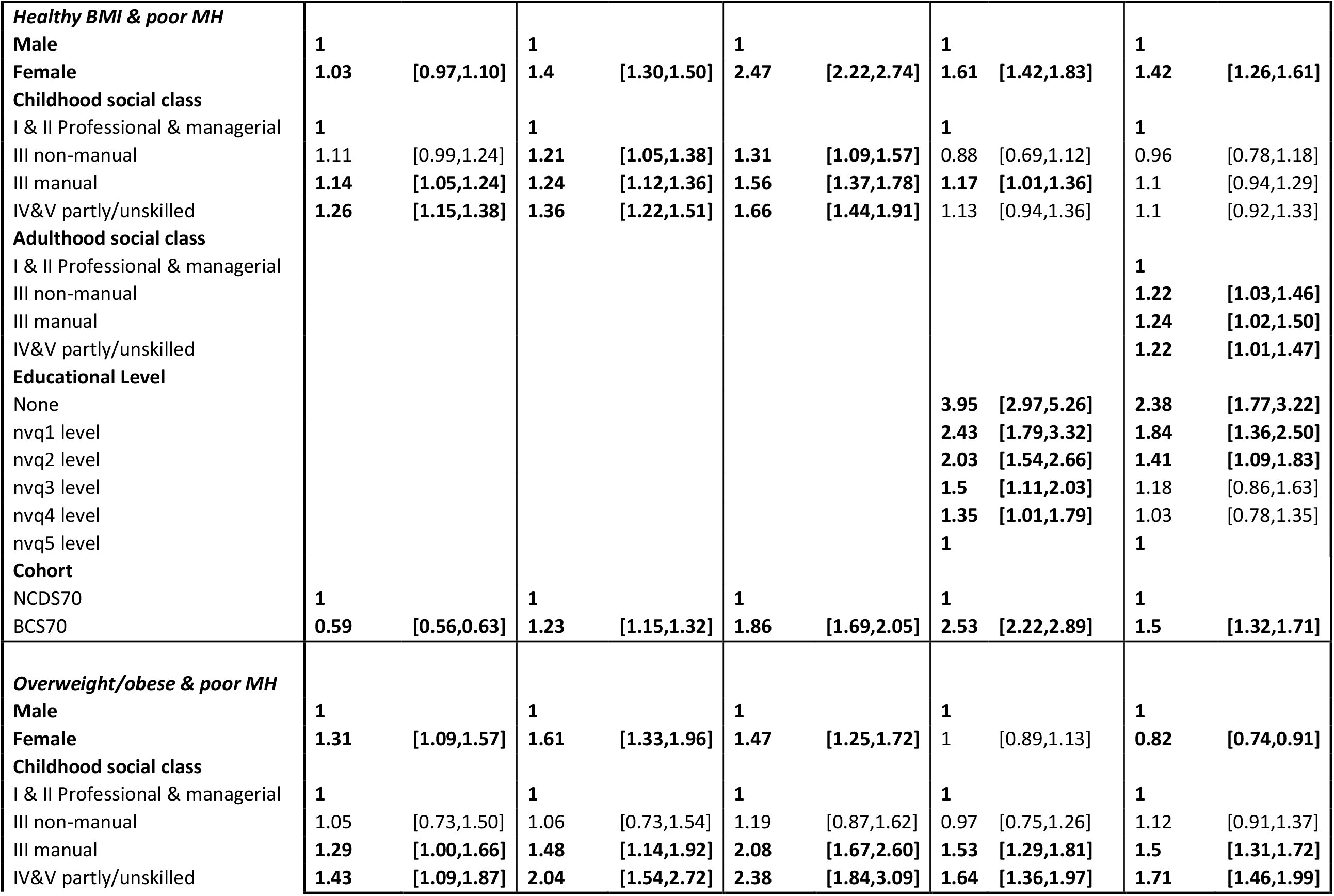
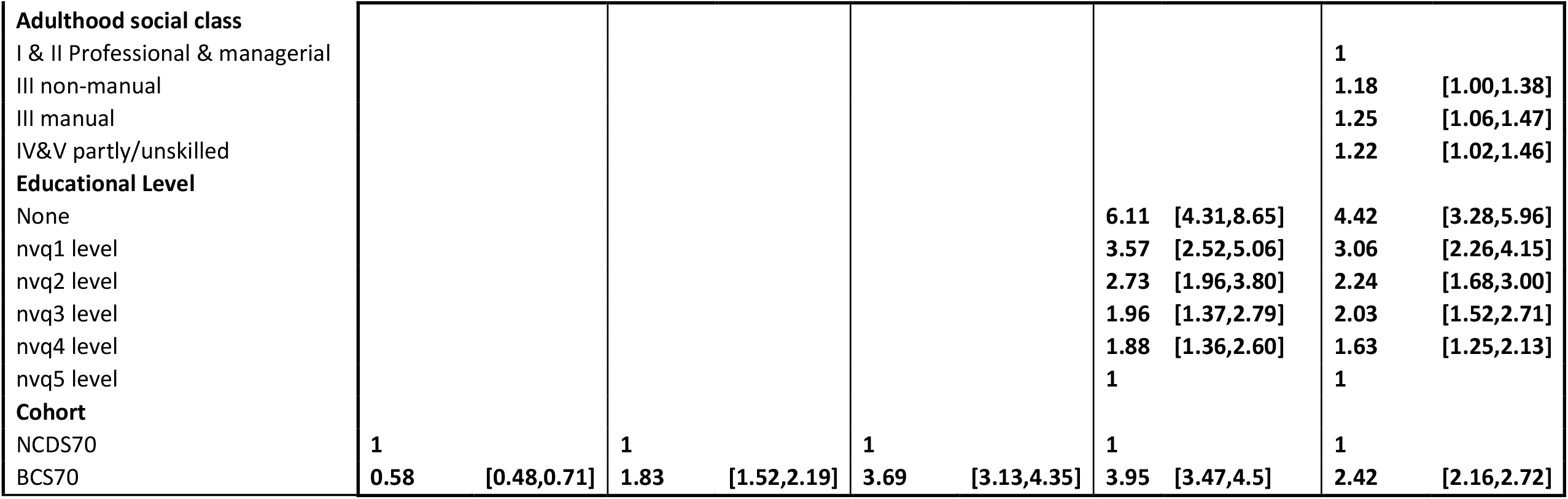
Relative risk ratios (RRR) for i. overweight or obesity and normal mental health, ii. Healthy BMI and poor mental health and iii. Overweight or obesity and poor mental health in 30 868 participants from the 1958 National Child Development Study and the 1970 British Cohort Study. Healthy BMI and normal mental health is the reference category.

**Supplemental Table 5.**
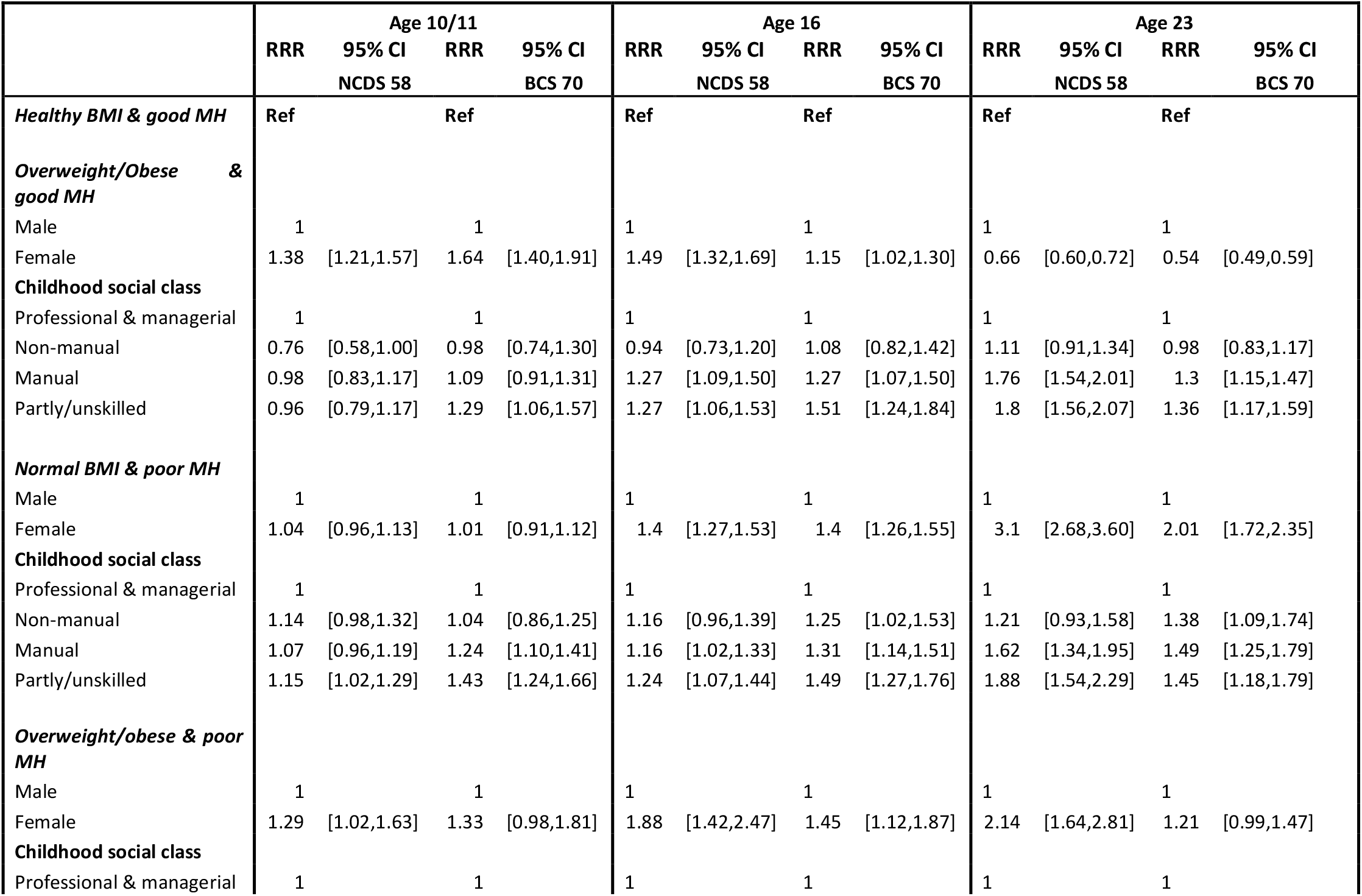
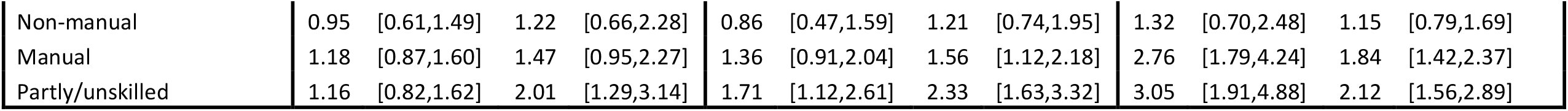
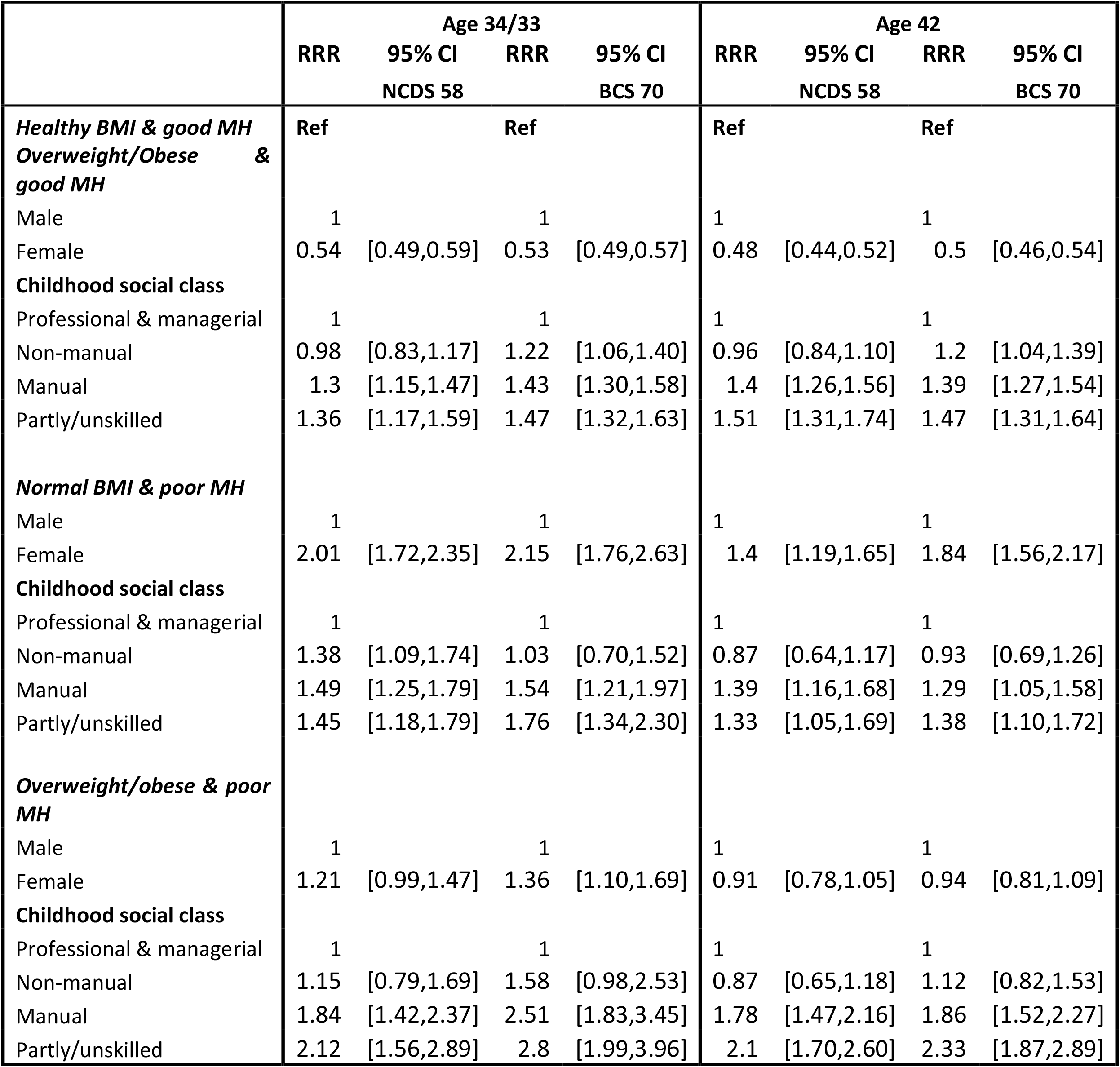
Relative risk ratios (RRR) between childhood social class and risk for i. overweight or obesity and normal mental health, ii. Healthy BMI and poor mental health and iii. Overweight or obesity and poor mental health in 30 868 participants from the 1958 National Child Development Study and the 1970 British Cohort Study. Healthy BMI and normal mental health is the reference category.

**Supplemental Figure 2 A, B, C & D.**
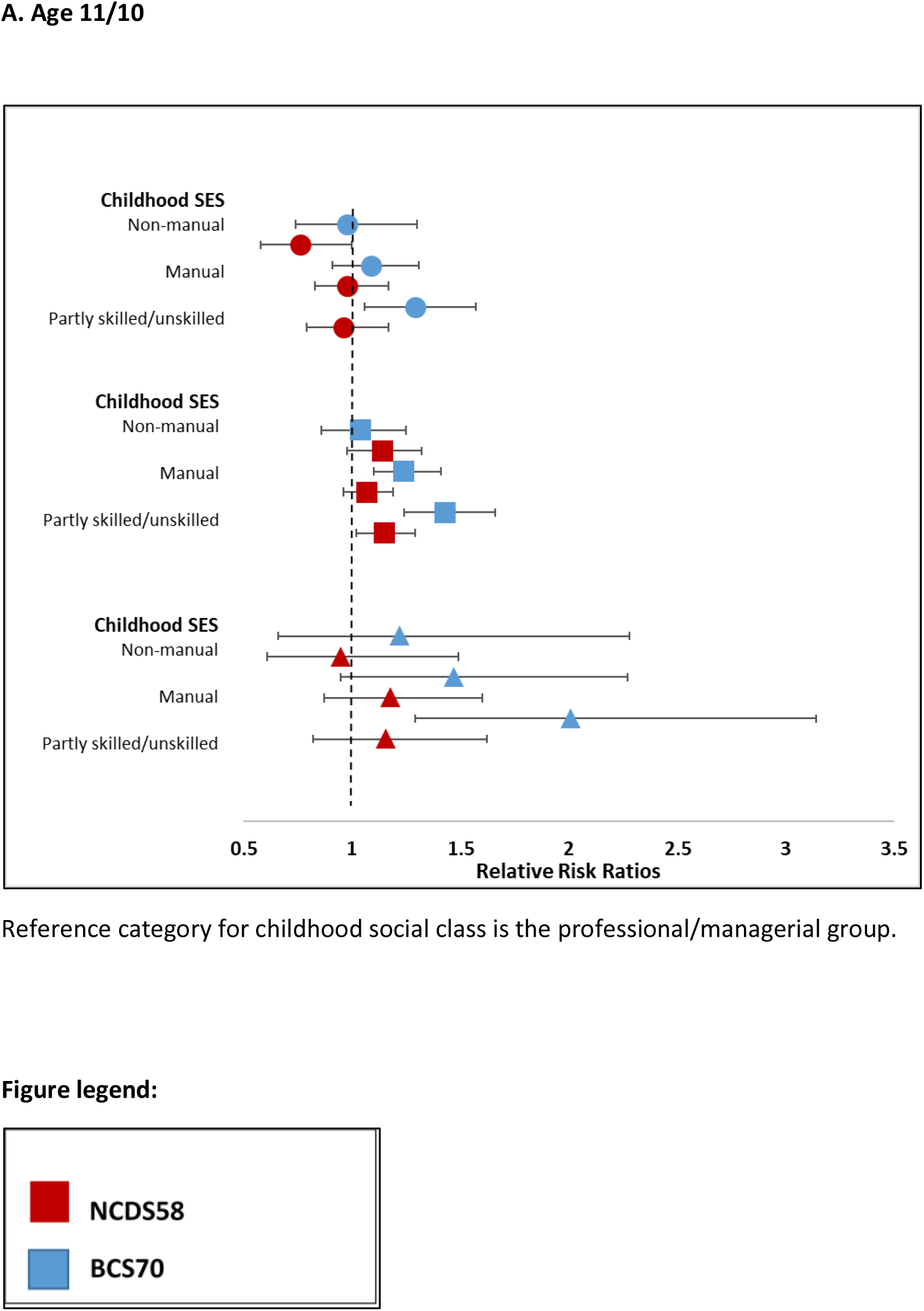
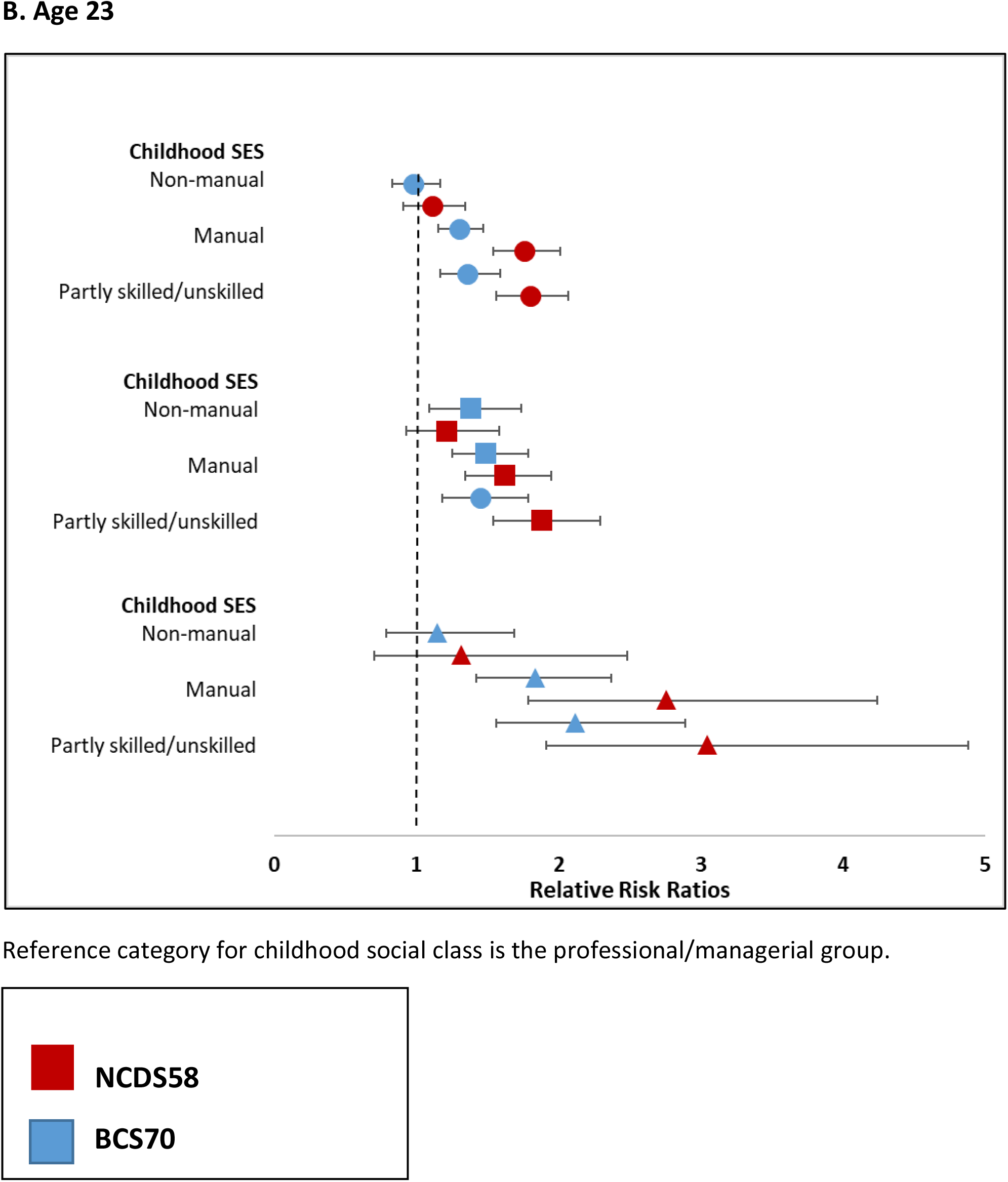
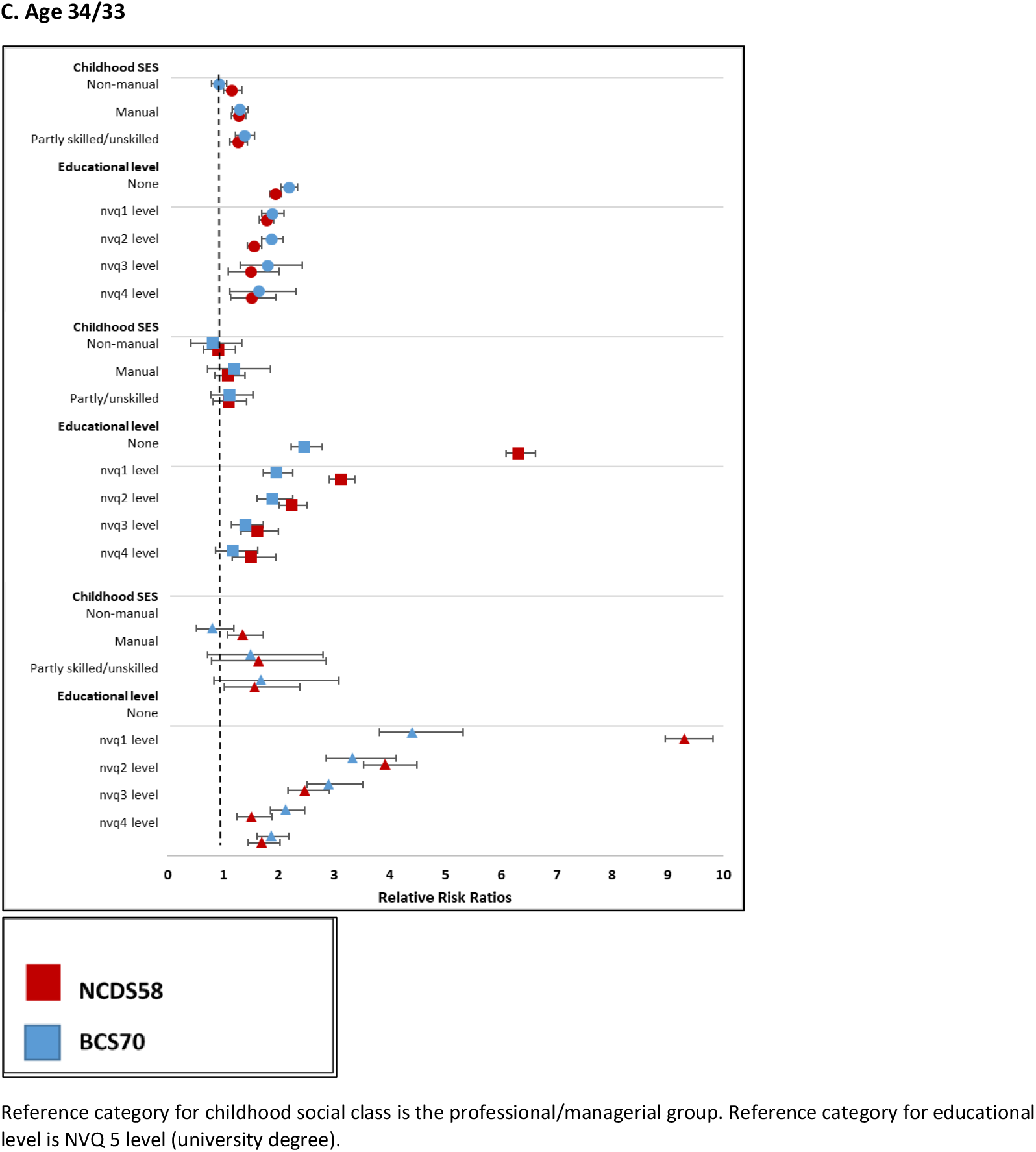
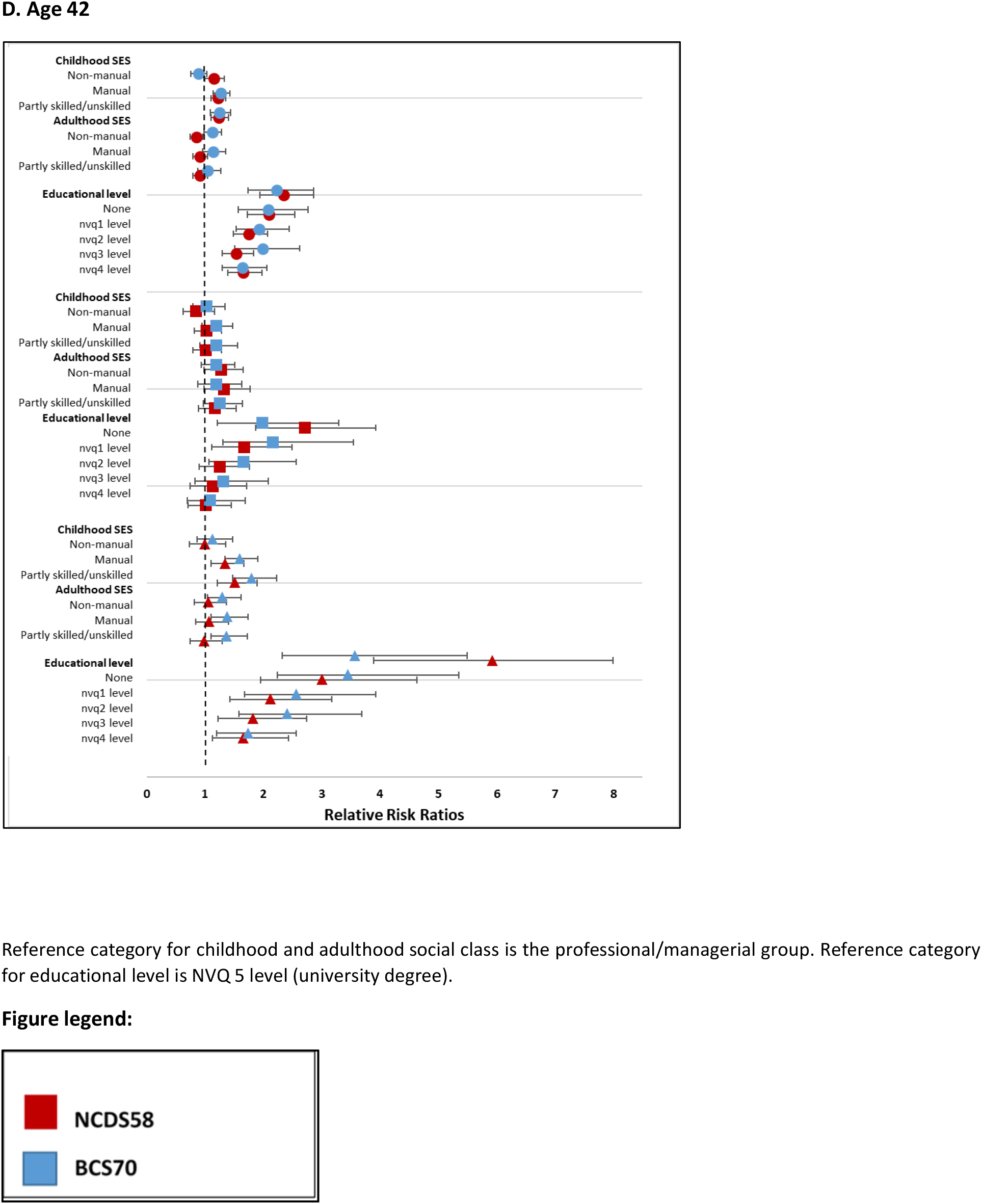
Relative risk ratios (RRR) for i. overweight or obesity and good mental health, ii. Healthy BMI and poor mental health and iii. Overweight or obesity and poor mental health in 30 868 participants from the 1958 National Child Development Study and the 1970 British Cohort Study, stratified by cohort. Healthy BMI and good mental health is the reference category

